# High magnitude of lung function decline among heroin users attending Medication-Assisted Therapy clinic at Muhimbili National Hospital, Tanzania

**DOI:** 10.1101/2025.01.01.25319867

**Authors:** Alex Nkongoki, Emmanuel Balandya, Simon Mamuya, Jessie Mbwambo

**Author notes:** Corresponding author Email. (AN). These authors contributed equally to this work (Alex Nkongoki, Emmanuel Balandya).

## Abstract

Heroin users have a high burden of respiratory morbidity, including premature lung function decline at age of 41 years. This early decline in lung function has been attributed to exposure to multiple risk factors. Further, methadone treatment has been found to aggravate lung function decline and independently cause asthma. However, the lung function status among heroin users on Medication-Assisted Therapy clinic in Tanzania is yet to be studied. This study aimed to assess the magnitude, pattern, and factors associated with lung function decline among heroin users attending MAT clinic at the Muhimbili National Hospital in Dar-es-salaam, Tanzania. This was a quantitative, analytical cross-sectional study conducted between May, 2022 – July, 2023. Individuals aged 18 years or above with heroin use disorder on the maintenance phase of treatment were recruited into the study. Data were collected using a questionnaire, and lung functions were measured using a portable spirometer. Lung function decline was defined as the percentage of participants with FEV1 <70%. The association between socio-demographic characteristics, substance use history and comorbidities with lung function decline was assessed via binary logistic regression. We enrolled 302 participants into the study (mean age of 42.78±7.56 years), whereby 95.7% were male. Of the study participants, 28.5% had lung function decline with mean age at onset of 44 **±**8 years. Restrictive lung disease was the commonest pattern of lung function decline with a proportion of 13.2%. Independent predictors of lung function decline were being underweight (aOR 4.73, 95% CI 2.61-8.59, p<0.001), living with HIV infection (aOR 2.61, 95% CI 1.20-5.66, p=0.016) and having a history of pulmonary TB (aOR 2.48, 95% CI 1.48-4.17, p=0.001). Heroin users on methadone therapy in Dar-es-salaam have high magnitude of lung function decline compared to the general population. Routine lung function testing is recommended.

## Introduction

Heroin users experience high and increasing burden of respiratory symptoms and decline in lung function that exceeds normal age-related decline observed among tobacco smokers with chronic obstructive lung disease (COPD) (1). Of note, the heroin users have been reported to have high rate of respiratory morbidity, hospital admission and a standardized mortality rate of 8.9% (2). This high burden of poor respiratory health has been attributed by exposure to multiple risk factors that include substance use and commodities such as HIV infection, pulmonary tuberculosis, and undernutrition.

Most heroin users attending Medication Assisted Therapy (MAT) clinics have the history of using more than one substance at certain point in their lifetime (polysubstance use)(3). These substances include tobacco, cannabis, heroin, cocaine, and ecstasy. Heroin can be used through injection, smoking, sniffing, and inhalation of fumes, commonly referred to as “*chasing the dragon*”. Injecting heroin has been associated with harmful effects such as bloodborne infections (HIV, hepatitis C, sepsis), septic and foreign body embolism and more severe symptoms of dependence(4). Furthermore, injecting heroin has been shown to cause lung fibrosis and reduce diffusion capacity of the lungs (5).

In recent years, heroin smoking rather than injecting has been used as a major means of harm reduction. Most heroin smokers use heroin combined with other substances (4). However smoking heroin is not without risk as lung function decline and overdose may occur. Inhaling heroin and other substances cause repeated airway exposure to irritants that can cause damage the airways and lungs (5). Airway exposure to toxic substances causes abnormal re-epithelialization and re-innervation of the bronchial mucosa that leads to airway hyperresponsiveness. Furthermore, heroin and cigarette smoking can cause release of histamine in the airway leading to pulmonary vein vasoconstriction, increased capillary permeability and bronchospasm (3). Heroin smoking can lead to the development of irreversible airflow obstruction hence lung function decline(6). Moreover, just like other opioids, heroin use can cause respiratory depression (3). Heroin users prescribed with methadone have high magnitude of asthma and COPD. Methadone is an independent risk factors for development of asthma(2).

Heroin users have high magnitude of HIV infection compared to the general population. In Tanzania, it is estimated that the prevalence of HIV infection among people who inject heroin to be 11% - 51.1%(7). This magnitude is higher compared to the 4.5% of the general population in Tanzania(8). People living with HIV have a rapid decline in Forced Vital Capacity (FVC) and Forced Expiratory Volume in 1 second (FEV1)(9). This simultaneous decline in FVC and FEV1 indicates the presence of Restrictive lung disease, and/or mixed abnormalities(9). Besides HIV infection, heroin users are also at risk of malnutrition due to poor food intake which can lead to underweight(10) and associated reduction in FEV1 and FVC (11). Furthermore, heroin users in Tanzania also have a higher magnitude of pulmonary tuberculosis (TB) of 56% compared to 2.6% in the general population(12). Pulmonary tuberculosis has been associated with low lung functions(13).

Lung function decline is the degenerative change that occurs with normal physiological aging. These changes become more marked after the age of 60 years (14). Studies show that FEV1 declines at a rate of 35mls/year after the age of 65years (14). Heroin users have premature lung function decline with mean age at onset of 41 years that exceeds age-related decline observed among tobacco smokers with COPD that normally starts to occur at the mean age of 56 to 63 years (11). Further, methadone that is used in the treatment of heroin used disorder has been show to aggravate lung function decline. A study done in Tanzania showed that there is a prevalence of 17% of spirometry lung function decline among people in rural areas (15). However, the extent of lung function decline among heroin users who receive methadone treatment in Tanzania has not been explored. In the absence of such insight, it is not possible to launch routine screening and treatment services for lung diseases in this population.

This study aimed to uncover the magnitude of lung function decline and its associated factors among heroin users on methadone therapy attending the MAT clinic at the Muhimbili National Hospital (MNH) in Dar-es-salaam, Tanzania with the goal to provide insights that will enable the provision of evidence-based recommendations for the implementation of routine screening and treatment of lung diseases in this patient population.

## Materials and Methods

### Study design

This was a quantitative, analytical cross-sectional study that was conducted among clients with heroin use disorder on methadone treatment attending MAT clinic at the Department of Psychiatry and Mental Health, Addiction Unit, MNH in Dar-es-salaam, Tanzania between May, 2022-July 2023.

### Sample size estimation

The sample size was calculated using fisher’s formula;

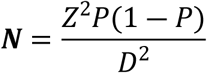

Where;

N-minimum sample size

Z-the standard deviation at 95% confidence interval (=1.96) P-the proportion of lung function decline (28%) (9).

D-level of precision (±0.05)

The minimum sample size was 310.

### Sampling technique

Systematic sampling was used to enroll participants into the study. This method was chosen because it was simple, time-saving and every client had an equal chance of being selected. A randomly ordered list of clients attending MAT clinic per day at MNH was obtained. Starting at a random point, we took every third client until 310 participants were enrolled. A fourth client was taken for the third client who was not available or defaulted from treatment.

### Inclusion and exclusion criteria

We included all heroin users above 18 years on maintenance phase of treatment. Clients with active pulmonary tuberculosis (TB) on intensive phase of treatment for TB and those on assisted therapy using suboxone or buprenorphine were excluded from the study.

### Variables

Dependent variable was forced expiratory volume in one second (FEV1). Independent variables were age, sex, body mass index (BMI), history of heroin smoking and injection, cigarette smoking, cannabis smoking, cocaine smoking, methadone use, HIV infection, and history of pulmonary TB.

### Data collection tools

Data were collected using a questionnaire which was administered to each study participant via one-on-one interview with the investigator. The questionnaire had five parts; the first part collected data on socio-demographic characteristics, the second part on lifestyle, the third part on the self-reported history of comorbidities, the fourth part on common respiratory symptoms and the last part recorded the lung function parameters following spirometry. A portable EasyOne^TM^ diagnostic spirometer model: 2001, SN:7294/2009 was used to measure lung functions. A weighing scale with a stadiometer was used to measure the weight and height of the participants, respectively. These data were collected between 20^th^ march 2023 up to 22^nd^ April 2023.

### Measurement of lung functions

Lung functions were measured using the spirometer with the participant standing. The participant was asked to maximally inhale with a rapid and sharp take off. After that, the participant was instructed to exhale rapidly and forcefully through the spirometer until the alarm of the spirometer went off, which occurred after at least 6 seconds. The participant was required to do three maneuvers. The result of each one maneuver was individually assessed based on the output on the spirometer which included categories A – excellent maneuver; B – very good, C – Good, D – Satisfactory and F – poor maneuver. Only categories A and B, “*good effort*”, were acceptable. In case of more than one acceptable effort, the best effort was recorded.

### Validity and reliability

The validity of the questionnaire was ensured through pre-testing of the questionnaire. Reproducibility and acceptability of the spirometry results were considered. We only included participants with African descent in the study.

### Data management

Data from completed questionnaires were coded and entered into the computer software SPPSS version 29. Categorical variables were summarized in frequencies and percentages, and continuous variables were summarized in means and standard deviations. The magnitude of lung function decline was calculated as the total number (frequency) and percentage of respondents with FEV1 <70%. The pattern of lung function decline was considered obstructive when FEV1/FVC ratio was <0.7, restrictive when FEV1/FVC was normal or elevated with FVC<80%, and mixed abnormality when FEV1/FVC ratio was <0.7 and FVC was <80%. The continuous variables (age, and BMI) were categorized into groups before analysis of association with lung function decline. The body mass index (BMI-kg/m^2^) was classified; <18.5 as underweight, 18.5-24.9 as normal weight, 25-29.9 as overweight and >30 as obesity. The relationship between sociodemographic characteristics, comorbidities, and substance use history with lung function decline was assessed using binary logistic regression with statistical significance determined via confidence interval at 95% with a p-value less than 0.05 taken as statistically significant.

## Results

### Socio-demographic, substance use history and clinical characteristics of the study participants

We enrolled 310 participants into the study. Out of these, 8 participants were excluded from the study because they could not complete the spirometry test. The mean age of the 302 participants who completed the questionnaire and spirometry was 42.78±7.56 years where 95.7% were male and 35.1% were single. The mean weight of these participants was 58.8±8.91 kg, mean height was 168.27±8.00 cm and the mean BMI was 20.82±2.93 kg/m^2^. The proportion of underweight participants was 20.9% (Table 1).

**Table 1:** Socio-demographic characteristics of the study participants.

| Variable | Parameter | Frequency (n) | Percent (%) |
| --- | --- | --- | --- |
| Age group (Years) | 23-32 | 33 | 10.9 |
|  | 33-42 | 104 | 34.4 |
|  | 43-52 | 136 | 45 |
|  | 53-63 | 29 | 9.6 |
| Sex | Male | 289 | 95.7 |
|  | Female | 13 | 4.3 |
| Marital status | Single | 106 | 35.1 |
|  | Married | 68 | 22.5 |
|  | Cohabiting | 72 | 23.8 |
|  | Divorced | 48 | 15.9 |
|  | widow/widower | 8 | 2.6 |
| Education level | No education | 20 | 6.6 |
|  | Primary | 65 | 21.5 |
|  | Secondary | 211 | 69.9 |
|  | College | 6 | 2 |
| Occupation | Unemployed | 31 | 10.3 |
|  | Self-employed | 249 | 82.5 |
|  | Employed | 22 | 7.3 |
| Housing | Homeless | 5 | 1.7 |
|  | Rented house | 102 | 33.8 |
|  | Own house | 27 | 8.9 |
|  | Family house | 168 | 55.6 |
| BMI (Kg/m<sup>2</sup>) | Normal weight | 214 | 70.9 |
|  | Underweight | 63 | 20.9 |
|  | Overweight | 25 | 8.3 |

Current cigarette smokers were 83.8% of study participants with an average of 8.62±5.10 cigarettes smoked per day. The mean smoking duration was 21±10 years. Current smokers were classified based on the magnitude of pack-year as 1-20 - light smokers, 20-40 - moderate smokers and >40 - heavy smokers. The mean pack-year was 9.53±8.35, with 90.1% being light smokers (Table 2).

**Table 2:** Substance use history of the study participants.

| Variable | Parameter | Frequency (n) | Percent (%) |
| --- | --- | --- | --- |
| Cigarette smoking status | Never smoked | 3 | 1 |
|  | Ex-smoker | 46 | 15.2 |
|  | Current smoker | 253 | 83.8 |
| Cigarettes smoked per day | <20 | 227 | 89.7 |
| | $\geq 20$ | 26 | 10.3 |
| Smoking duration (years) | $\leq 20$ | 114 | 45.1 |
|  | >20 | 139 | 54.9 |
| Smoking intensity (pack-years) | Light smoker | 228 | 90.1 |
|  | Moderate smoker | 25 | 9.9 |
| Heroin sachets smoked per day | <20 | 259 | 85.8 |
| | $\geq 20$ | 43 | 14.2 |
| Heroin use duration (years) | <20 | 195 | 64.6 |
| | $\geq 20$ | 107 | 35.4 |
| History of injecting heroin | Yes | 60 | 19.9 |
|  | No | 242 | 80.1 |
| Cannabis use status | Never smoked | 41 | 13.6 |
|  | Ex-smoker | 261 | 86.4 |
| Cocaine use status | Never smoker | 217 | 71.9 |
|  | Ex-smoker | 85 | 28.1 |
| Methadone dose (mg) | <80 | 81 | 26.8 |
|  | 80-120 | 102 | 33.8 |
|  | >120 | 119 | 39.4 |
| Duration on methadone (years) | <5 | 217 | 71.9 |
|  | >5 | 85 | 28.1 |

The participants averaged 9.88±7.13 sachets of heroin smoked per day, with a mean duration of heroin smoking of 15.33±7.79 years. Additionally, 19.9% of the participants had history of injecting heroin. Regarding MAT, the average dose of methadone was 131±88 mg and the mean duration the participants were on methadone treatment was 4.23±2.62 years. Furthermore, 86.4% of participants were ex-cannabis smokers and 28.1% of participants were ex-cocaine smokers (Table 2).

The magnitude of participants who self-reported being asthmatic was 8.3%, while the proportion of participants living with HIV infection on treatment was found to be 9.6%. Furthermore, 37.7% of the participants reported experiencing at least one respiratory symptom over the past three months, with 22.2% specifically reporting cough (Table 3).

**Table 3:**
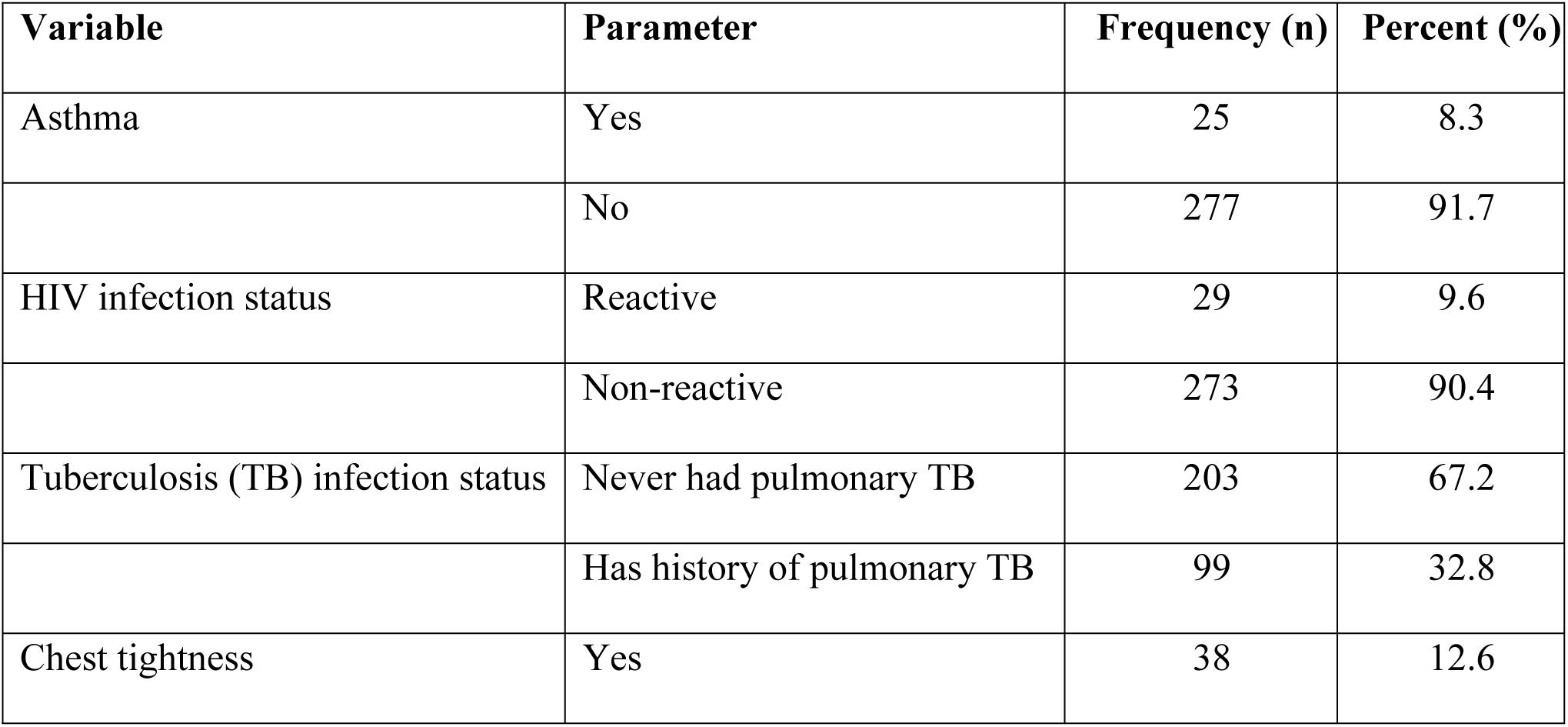
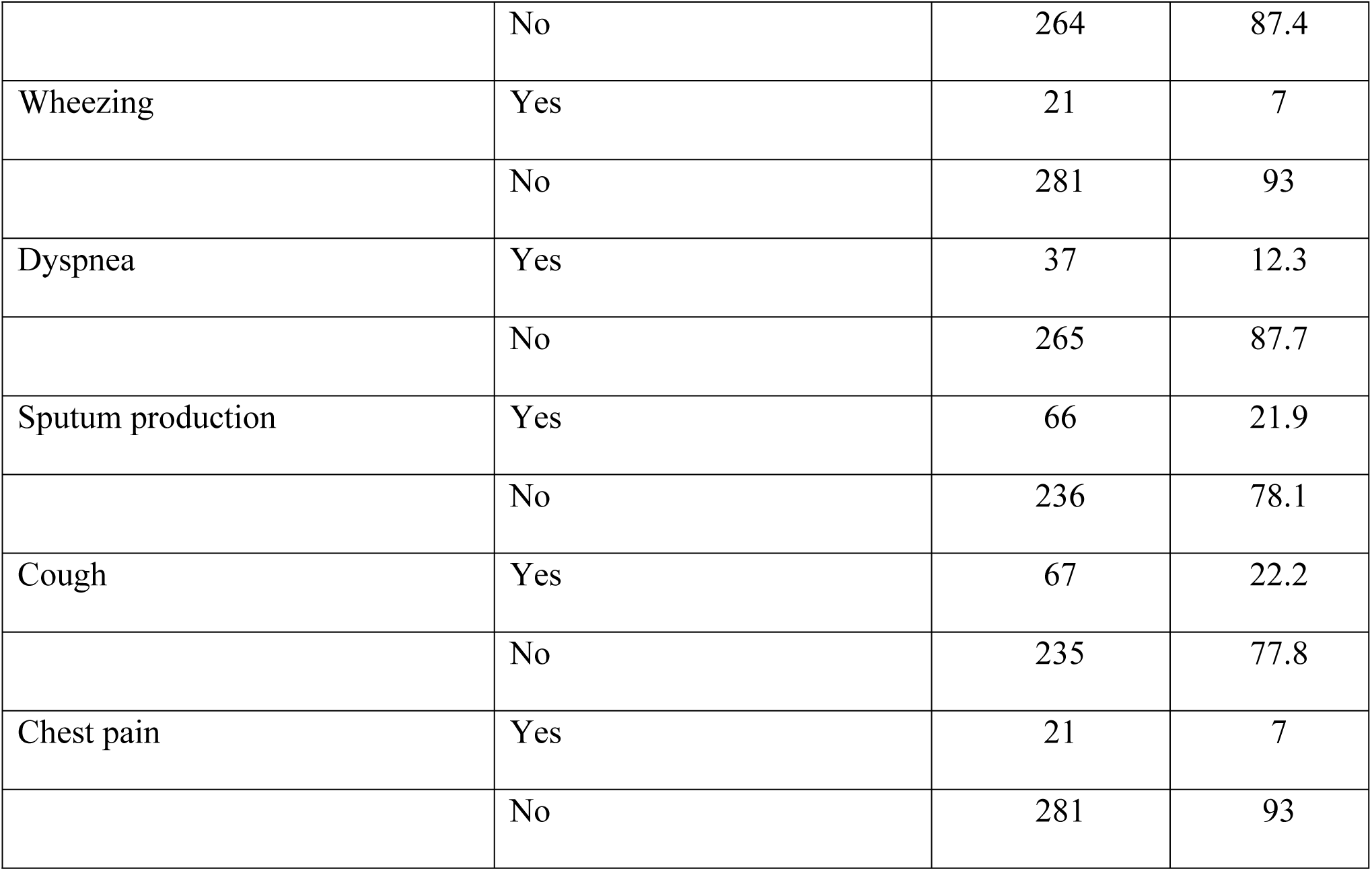
Clinical Characteristics of the Study Participants.

| Variable | Parameter | Frequency (n) | Percent (%) |
| --- | --- | --- | --- |
| Asthma | Yes | 25 | 8.3 |
|  | No | 277 | 91.7 |
| HIV infection status | Reactive | 29 | 9.6 |
|  | Non-reactive | 273 | 90.4 |
| Tuberculosis (TB) infection status | Never had pulmonary TB | 203 | 67.2 |
|  | Has history of pulmonary TB | 99 | 32.8 |
| Chest tightness | Yes | 38 | 12.6 |
|  | No | 264 | 87.4 |
| Wheezing | Yes | 21 | 7 |
|  | No | 281 | 93 |
| Dyspnea | Yes | 37 | 12.3 |
|  | No | 265 | 87.7 |
| Sputum production | Yes | 66 | 21.9 |
|  | No | 236 | 78.1 |
| Cough | Yes | 67 | 22.2 |
|  | No | 235 | 77.8 |
| Chest pain | Yes | 21 | 7 |
|  | No | 281 | 93 |

### Lung function test results of the study participants

The proportion of participants with lung function decline was 28.5% with a mean FEV1 of 2.42±0.72 L (Table 4). Moreover, the proportion of participants with possible obstructive lung disease was 11%, while restrictive lung disease accounted for 13.2%, and mixed abnormality (obstructive and restrictive lung disease) was observed in 4.3% of participants (Table 5). The mean age of onset of lung function decline was found to be 44±8 years.

**Table 4:** Lung Function results of the study participants.

| Variables | Parameter | Mean | SD |
| --- | --- | --- | --- |
| Forced vital capacity | FVC (L) | 3.66 | 1.12 |
|  | FVC (% predicted) | 97.68 | 30.14 |
| Forced expiratory volume in 1 second | FEV1 (L) | 2.42 | 0.72 |
|  | FEV1 (% predicted) | 78.88 | 22.46 |
| FEV1/FVC | FEV1/FVC (L) | 67.41 | 15.64 |
|  | FEV1/FVC (% predicted) | 82.56 | 19.07 |
| Peak expiratory flow rate | PEFR (mls/s) | 336.44 | 133.18 |
|  | PEFR (% predicted) | 69.92 | 32.08 |
| Forced expiratory flow 25-75 | FEF 25-75 (L) | 1.75 | 1.14 |
|  | FEF25-75 (% predicted) | 52.67 | 33.18 |

**Table 5:** Magnitude and pattern of lung function decline.

| **Variable** | **Parameter** | **Number (n)** | **Percent (%)** |
| --- | --- | --- | --- |
| Lung function status | Lung function decline | 86 | 28.5 |
|  | Normal lung function | 216 | 71.5 |
| Pattern of lung function decline | Obstructive lung disease | 33 | 11 |
|  | Restrictive lung disease | 40 | 13.2 |
|  | mixed abnormality | 13 | 4.3 |
|  | Normal lung function | 182 | 60.3 |

### Risk factors associated with lung function decline

Compared to individuals who had normal body weight, participants who were underweight were 4.7 time more likely to have lung function decline (aOR 4.73, 95% CI 2.61-8.59, p<0.001). (Table 6).

**Table 6:** Association of sociodemographic factors with lung function decline.

| Variable | Parameter | *LFD n (%) | **NLF n (%) | aOR | 95% CI | p-value |
| --- | --- | --- | --- | --- | --- | --- |
| Age group (years) | 23-32 | 9 (27.3) | 24 (72.7) | 1 | reference |  |
|  | 33-42 | 24 (23.1) | 80 (76.9) | 0.80 | 0.33-1.95 | 0.624 |
|  | 43-52 | 43 (31.6) | 93 (68.4) | 1.23 | 0.53-2.88 | 0.628 |
|  | 53-63 | 10 (34.5) | 19 (65.5) | 1.40 | 0.48-4.15 | 0.540 |
| Sex | Female | 5 (38.5) | 8 (61.5) | 1 | Reference |  |
|  | Male | 81 (28.0) | 208 (72) | 0.62 | 0.19-1.96 | 0.419 |
| BMI (kg/m<sup>2</sup>) | Normal weight | 36 (57.1) | 27 (42.9) | 1 | reference |  |
|  | Underweight | 47 (22.0) | 167 (78) | 4.73 | 2.61-8.59 | <0.001 |
|  | Overweight | 3 (12.0) | 22 (88.0) | 0.48 | 0.14-1.69 | 0.255 |

\*Lung function decline \*\*Normal lung function aOR- adjusted odds ratio CI-confidence interval

Furthermore, participants living with HIV infection were 2.6 times more likely to have lung function decline compared to those who were HIV negative (aOR 2.61, 95% CI 1.20-5.66, p = 0.016) (Table 7). Similarly, participants with history of pulmonary TB were 2.4 times more likely to have lung function decline compared to those with no history of pulmonary TB (aOR 2.48, 95% CI 1.48-4.17, p=0.001) (Table 7).

**Table 7:**
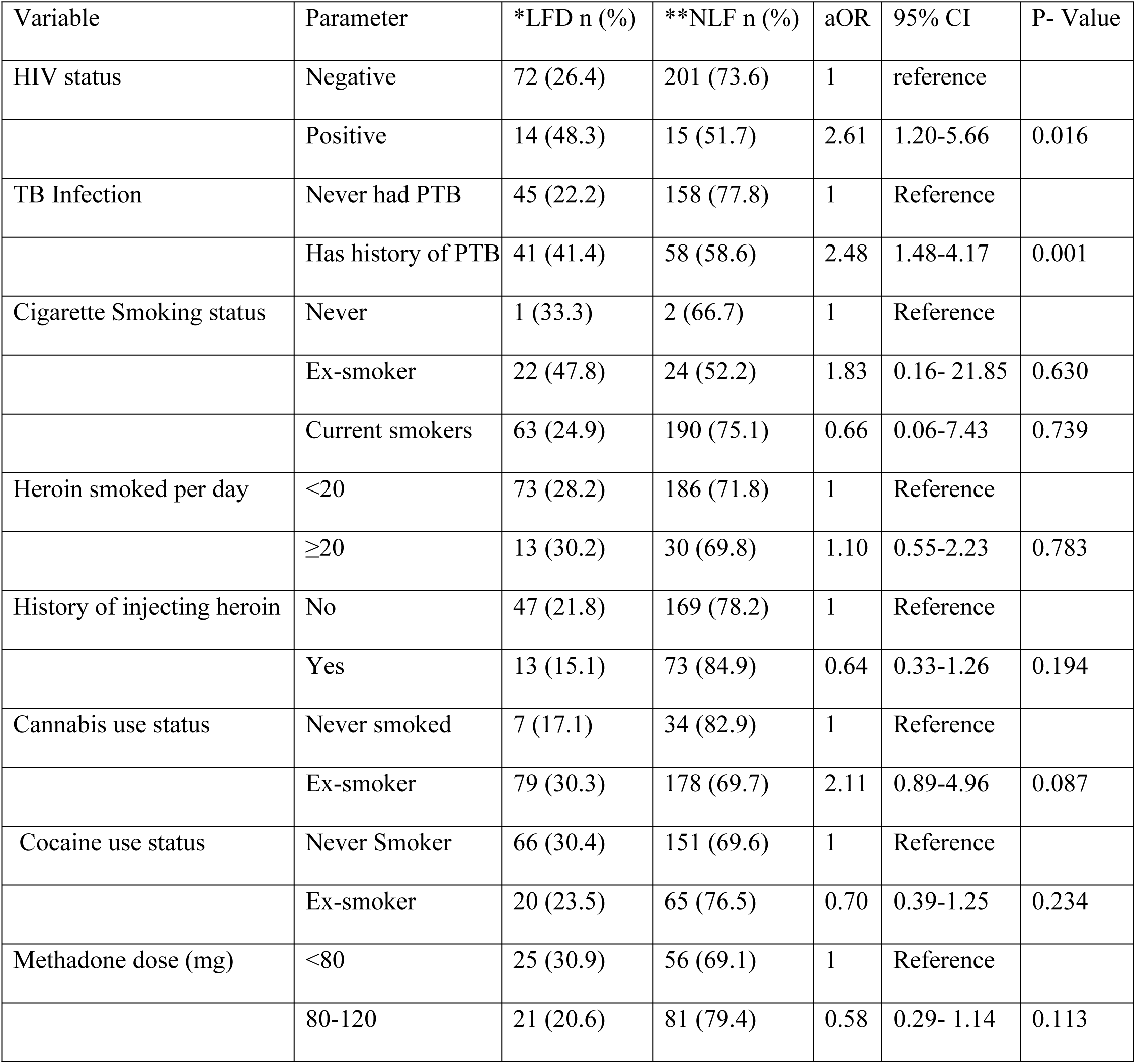
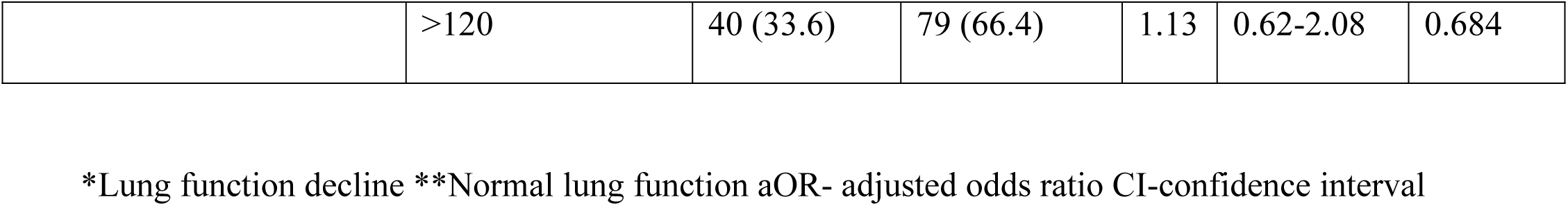
Association of clinical characteristics, substance use history with lung function decline.

## Discussion

Heroin users have high burden of respiratory morbidity, including premature decline in lung functions. Here, we show for the first time that over a quarter of heroin users who attend the largest MAT clinic in Tanzania have lung function decline. Further, we show that being underweight, living with HIV infection and having a history of pulmonary tuberculosis were independent positive predictors of lung function decline. This data calls for routine screening for lung function at the MAT across Tanzania and other countries with similar settings.

In our study, the mean age at onset of lung function decline was 44 years. These results are comparable with another study that reported 43 years as the mean age at onset of lung function decline(16), but is relatively younger compared to cigarette smokers who were reported to start having significant lung function decline at the age of 56 years(17). The proportion of lung function decline in our study population was 28.5%. This magnitude is higher compared to the proportion of lung function decline among cigarette smokers which is only 15% (5), and is likely due to other comorbid conditions that aggravate the lung functions among heroin smokers.

Restrictive lung disease was the commonest pattern of lung function decline among heroin users with a proportion of 13.2%. This dominance of restrictive lung disease is different from another study which reported 28% to have obstructive lung disease and 7% restrictive lung disease (16). The low level of self-reported cases of asthma (8.3%) further corroborates the study findings of lower proportion of obstructive lung disease (11%) in our study population. Despite these findings, no participant was diagnosed with any chronic airway disease prior participation in this study.

Underweight was found to be a risk a factor for lung function decline. These findings were similar to another study which showed that being underweight was associated with the decline in FEV1(11). Heroin smokers have poor appetite and poor food consumption and majority have protein-energy malnutrition that leads to underweight(10). As an opiate, heroin also causes anorexia and decreases food consumption, leading to under-nutrition (18). Studies show that, the rate of FEV1 decline is higher among people with low BMI (19).

HIV Infection was found to be another risk factor for development of lung function decline. Studies have shown that, the rate of FEV1 decline among people living with HIV was 10.5 mls/year. This decline is independent of smoking and might be attributed to the ongoing interstitial or small airway damage(9). Our findings are similar to other studies that showed HIV infection to be an independent risk factor for the development of COPD(20). Furthermore, people living with HIV have increased risk of non-infectious pulmonary diseases such as emphysema, pulmonary arterial hypertension and lung fibrosis(20).

In our study, 28.8% of the participants had past history of pulmonary TB. This magnitude is higher compared to 2.6% in the general population in Tanzania (12). Having a history of pulmonary TB was a risk factor associated with lung function decline. This finding can be explained by the healing by fibrosis and cavitation caused by TB infection. Pulmonary TB has been shown to cause airflow obstruction and lung function decline (13). A systematic review showed that patients with history of pulmonary tuberculosis experience respiratory morbidity post-treatment with 59% having abnormal lung functions: 22% obstruction, 23% restrictive and 15% mixed pattern (21). Further, studies have shown that restrictive lung disease is the commonest pattern of lung function decline in this population(22).

Despite the strength of being the first study in Tanzania to delineate the magnitude and determinants of lung function decline among heroin users attending a MAT, our study had a limitation of not being able to perform the bronchodilator reversibility test to differentiate asthma from COPD among patients with obstructive lung disease.

## Conclusion

Over a quarter of clients attending the MAT clinic had lung function decline. Being underweight, living with HIV infection and having a history of pulmonary TB were major risk factors for lung function decline in this population. Efforts should be made to include routine screening of lung function in order to address the undiagnosed and underdiagnosed lung diseases in this patient population.

## Ethical considerations

The approval to conduct this research was sought from the Muhimbili University of Health and Allied Sciences (MUHAS) Institutional Review Board (IRB) before the commencement of the study, and the permission to conduct the study at MNH was obtained from the hospital administration. Written informed consent was sought from the participants prior participation in the study. The participation was voluntary and confidentiality was ensured by interviewing one participant at a time in a private room. No identifying information was collected from the study participants. The investigators declare no conflicts of interest.

## Data Availability

All data are available from the the zenodo database. Can be accessed through DOI 10.5281/zenodo.14564232.

## Acknowledgment

We extend our sincere thanks to all the study participants, Ms Carol Lee for assistance in the acquisition of the disposable mouthpieces for the spirometer, Dr. Iddy Haruna from the Department of Psychiatry and Mental health at MNH for facilitating data collection, and to Mr. Seif Ngilini, Mr. Haruna Jumanne and Mr Alizeid Ramadhani (posthumous) for helping with the recruitment of the participants into our study.

## Notes

### Competing Interest Statement

The authors have declared no competing interest.

### Funding Statement

The author(s) received no specific funding for this work.

### Author Declarations

The approval to conduct this research was sought from the Muhimbili University of Health and Allied Sciences (MUHAS) Institutional Review Board (IRB). The approval number from the IRB was Ref. No. DA.25/111/01D/81.

